# Dispersion of a new coronavirus SARS-CoV-2 by airlines in 2020: Temporal estimates of the outbreak in Mexico

**DOI:** 10.1101/2020.03.24.20042168

**Authors:** G Cruz-Pacheco, J.F Bustamante-Castañeda, J.G Caputo, ME Jiménez-Corona, S Ponce-de-León

## Abstract

On January 23, 2020, China imposed a quarantine on the city of Wuhan to contain the SARS-CoV-2 outbreak. Regardless of this measure the new infection has spread to several countries around the world. Here, we developed a method to study the dissemination of this infection by the airline routes and we give estimations of the time of arrival of the outbreaks to the different cities.

In this work we show an analysis of the dispersion of this infection to other cities by airlines based on the classic model the Kermack and McKendrick complemented with diffusion on a graph composed of nodes which represent the cities and edges which represent the airline routes. We do several numerical simulations to estimate the date of arrival to different cities starting the infection at Wuhan, China and to show the robustness of the estimation respect to changes in the epidemiological parameters and to changes on the graph. we use Mexico City as an example. In this case, our estimate of the arrival time is between March 20 and March 30, 2020. This analysis is limited to the analysis of dispersion by airlines, so this estimate should be taken as an overestimate since the infection can arrive by other means.

This model estimates the arrival of the infectious outbreak to Mexico between March 20 and March 30. This estimation gives a time period to implement and strengthen preventive measures aimed at the general population, as well as to strengthen hospital infrastructure and training of human resources in health.

## Introduction

On December 31, 2019, China reported to the country office of the World Health Organization (WHO) an outbreak of cases of pneumonia of unknown etiology; From December 31 to January 3, 2020, 44 cases of pneumonia of unknown origin in Wuhan city, Hubei Province, were reported to WHO. On January 7, the Chinese authorities announced that it is a new coronavirus (nCoV-2019), now called SARS- CoV-2.^2^ Phylogenetic analyses were immediately performed showing that the new coronavirus is closer to the coronavirus SARS that emerged in Guandong, China in 2002; that with the MERS-CoV that emerged in Saudi Arabia in 2013.^2,3^ On January 30, the new coronavirus was declared the sixth Public Health Emergency of International Importance (PHEIC) according to the guidelines of the International Health Regulations.^4^ Until March 3, WHO reports 90,870 confirmed cases globally and 3,112 deaths, for China reports 80,304 confirmed cases and 2,946 deaths; outside of China 10,566 confirmed cases and, 166 deaths in 72 countries.^5^

According to the experience of the influenza pandemic due to influenza virus A(H1N1) in Mexico, 2009, it is cardinal to estimate the arrival time of the outbreak and spreading of SARS-CoV-2 to our country, in order to strengthen the activities according to the Health Emergency Preparedness and Response Plan, for the current SARS-CoV-2 epidemic that is evolving and from which new information is generated each day. For this reason new methods and techniques have been developed to qualitatively understand this type of phenomenon.

## Methods

In 1927 doctors Kermack and McKendrick developed a mathematical model to the study the Spanish Influenza pandemic of 1921.^6^ This model, with some modifications, was used for forecasts and analysis of the 2009 influenza outbreak in Mexico City.^7^ This same model, supplemented with classical diffusion (which in the context of an epidemic is managed as a dispersion of the infection) has been quite useful in studies of the spread of some infections as, for example, rabies.^8^ In this work we develop a new method to analyze the dissemination of a new virus as SARS-CoV-2 between different cities by airline routes with the purpose of estimating the arrival from one city to another. Our method uses the Kermack and Mckendrick model to simulate the way the outbreak grows and evolves when it arrives to the different cities and, also we use classical diffusion on a graph, to model the way the infection travels between cities by the airline routes.^9^ The purpose of using a simple model for our analysis is to have the least number of parameters, but with sufficient precision to be able to give an estimate of the date of arrival of the outbreak for example, to Mexico City. By this date, it is well established that infection by this type of coronavirus is transmitted person to person through drops expelled by an infected person when coughing or sneezing, it is also possible to be transmitted by contact with surfaces or objects contaminated with the virus and then one puts his hands to his mouth, eyes or nose.^10^ There is evidence that once the infectious outbreak has started there is some homogeneity in its development in the affected city for these influenza-like infections. For these reasons, the Kermack and McKendrick model is applicable in the cities that we use as nodes of the graph in which the edges represent the flights between one city and another. Brockmann and Helbling,^1^ used a similar model to study the dispersion of a general infection except that their transfer term is not a symmetric diffusion.

The basic reproductive number was estimated using the data reported for Hubei and Guangdong,^11^ an adjustment was made to the data at the beginning of the epidemic outbreak to estimate the force of infection λ, which is related to the basic reproductive number as follows R_0_ = 1 + λ / β. The setting of λ for Hubei and for Guangdong gives λ = 0.3. These estimates give an R_0_ = 2.5, which is in very good agreement with the estimate of Joseph T Wu^12^ of R0 = 2.53 and, is well into the interval of the estimations reported for JM Read.^13^ The diffusion coefficient D was estimated using numerical simulations of the model and the fact that a new outbreak started in Singapore around February 15, 2020. This gives a diffusion coefficient D = 10^−6^. In Figure 1 the nodes of the graph represent cities that were chosen because their airports function as important distribution points to other airports. In the model, some of these nodes also function as a representation of other airports in the region, for example, the Paris node represents the main international airports of that European region: Paris, Amsterdam and, Frankfurt. Using this graph and the estimated epidemiological parameters, the model was solved numerically to study in several scenarios the time of arrival of the infection to Mexico City.

**Figure 1.**
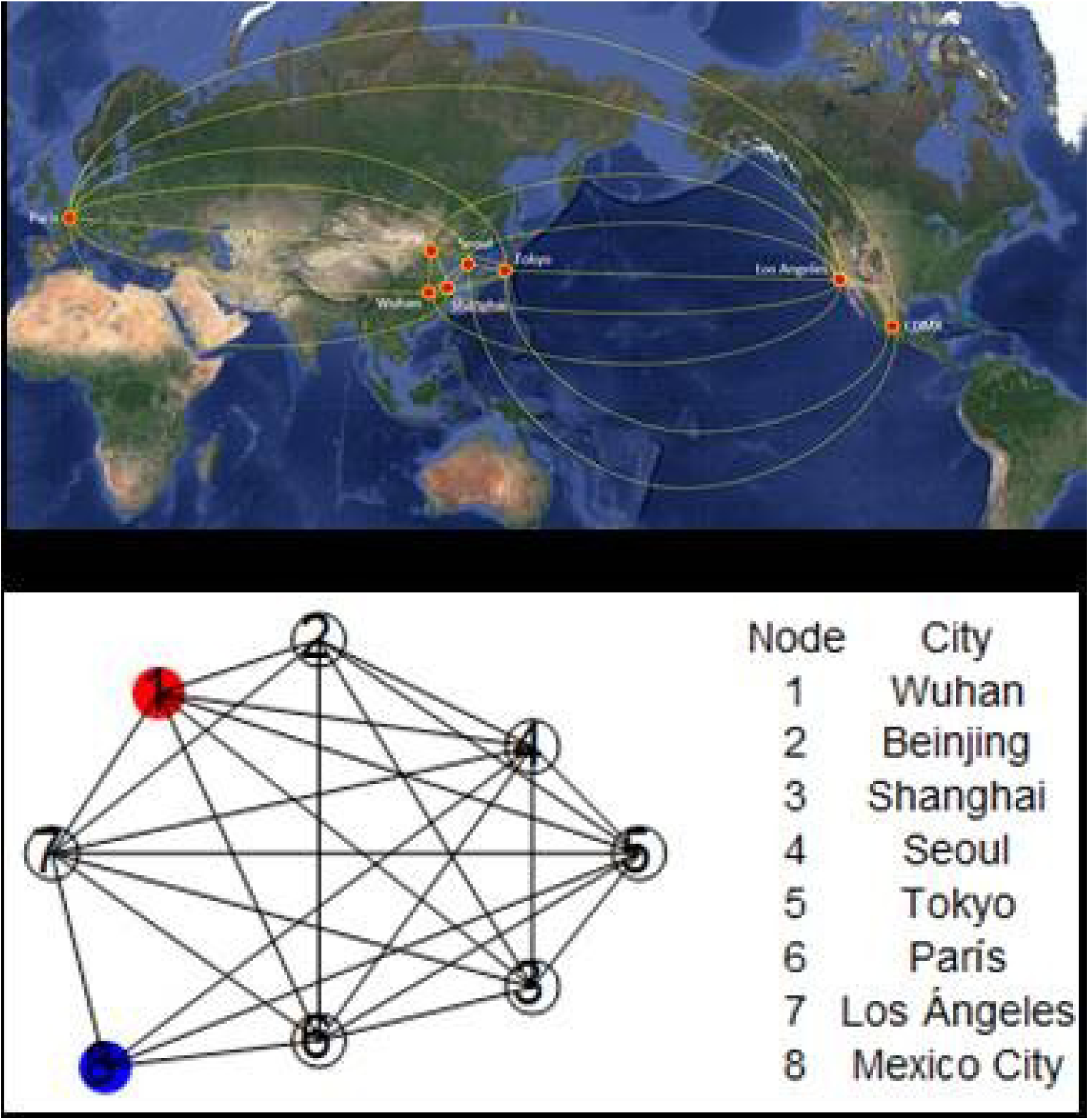
Cities considered as nodes in this analysis, as well as the corresponding graph.

## Results

Using the parameters estimated the model was solved numerically to study in several scenarios the arrival of the outbreak to Mexico City. All our simulations have initial conditions in which only node 1 corresponding to the city of Wuhan has been infected and the other cities are not yet infected or represent a very small number with respect to its population. In the city of Wuhan, an infected number of 20 is assumed,^14^ the initial date for this analysis was taken on January 10, 2020, as the starting point of the numerical simulations.

We solved numerically this model, the result is shown in figure 2, where the vertical axis is the infected proportion of the whole population of the city in question. The maximum of the first outbreak in red corresponding to Wuhan is reached around February 24, 2020, the peaks corresponding to nodes 2 through 7 are all one above the other, peak in violet, because the corresponding graph is all interconnected (they form what is called complete graph) if one excludes node 8, the remaining graph does not distinguish one node from the others. This numerical simulation also indicates that the maximum of the first curve reaches 0.23, this means that, at the maximum point of the graph, 23 percent of the urban area of Wuhan is infected. Data from Johns Hopkins University show that the number of reported cases is peaking at approximately 63,000. The urban area of Wuhan is around 20 million inhabitants (this is the population that serves Wuhan airport), 23% corresponds to 4,600,000, this indicates that for each case reported there are about 73 infected in the city. In this scenario, the infectious outbreak of node 8, peak in green, corresponding to Mexico City begins to grow between t=70 and t=80, which gives an arrival of the infectious outbreak to Mexico between March 20 and 30, blue vertical line. This information was made available to the Mexican Secretary of Health on February 24, 2020.

**Figure 2.**
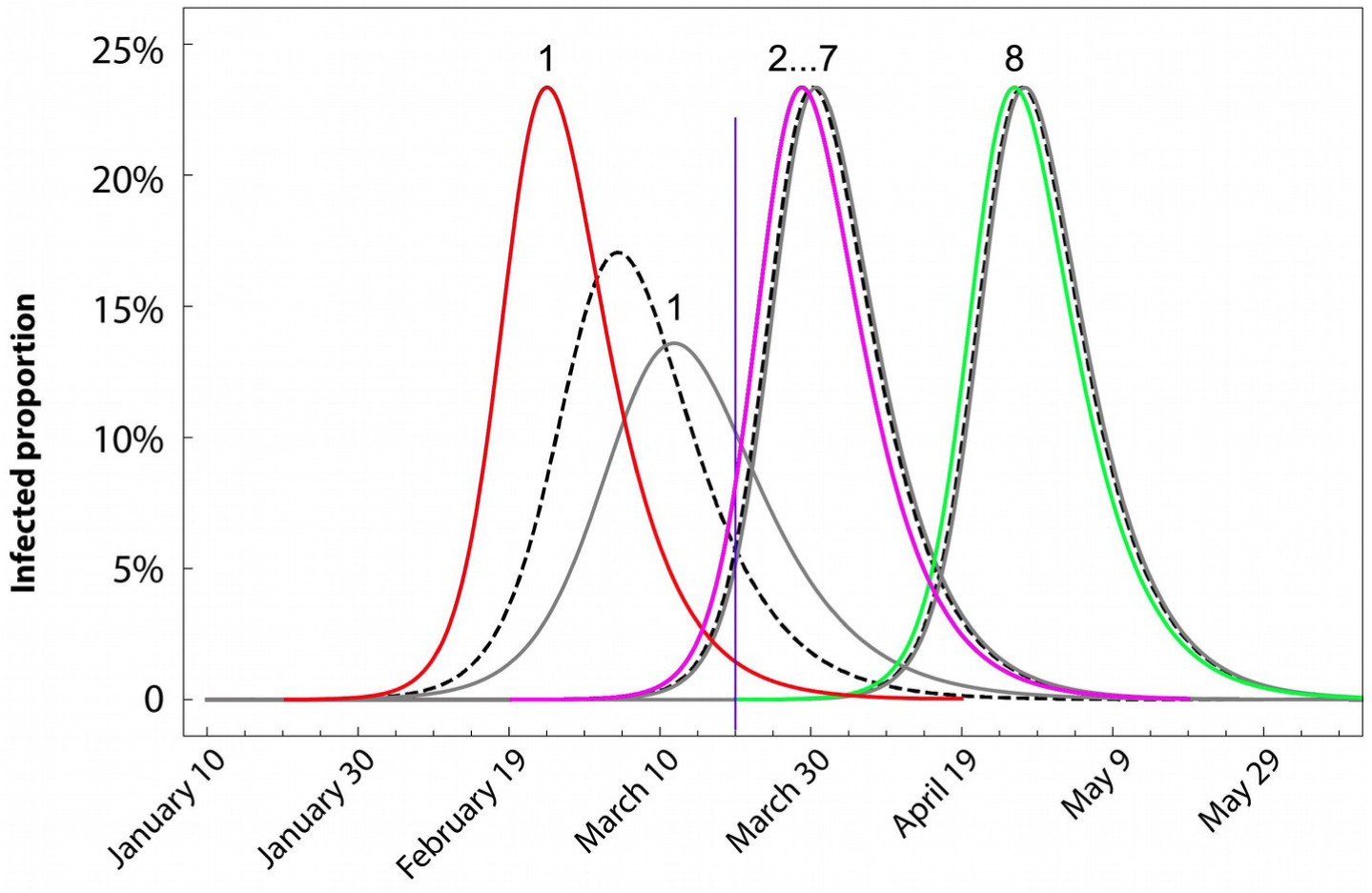
The curves in colors show a numerical simulation of the time evolution of an infectious outbreak that begins at node 1, Wuhan, red curve. After peaking at node 1, it spreads to nodes 2 to 7, violet curve. Finally, it arrives to node 8, Mexico City, green curve. The curves in black show a simulation in which R_0_ has been changed as follows: R_0_ = 2.1 (black in dashed line) and 1.9 (black). In both cases, the corresponding peaks at node 8 only gets slightly retarded.

To simulate the isolation measures imposed on Wuhan since the end of January, numerical simulations where performed with lowering the value of R_0_ from 2.5 to 2.1 first and then to 1.9, This is shown in figure 2 by the black dashed curve for R_0_ = 2.1 and by the black curve for R_0_ = 1.9. As can be seen, this does not affect the high and width of the rest of the peaks and only delays the arrival of the rest of the nodes including node 8 of Mexico City by around of 4 days. This shows that changes not yet so small in R_0_ do not substantially alter the result on the arrival of the outbreak at node 8. Simulations with R_0_ = 3, when they era adjusted to match the outbreak in Wuhan also only anticipated the outbreak in Mexico City by 4 days. We performed other numerical simulations changing the R_0_ in other nodes of the graph around 10% of its value, this changed the outbreaks at these nodes, but it did not change significantly, the date of arrival of the outbreak in Mexico City. Changes on the graph structure cutting some edges, meaning cancelling some airline routes, also did not modify significantly the date of arrival of the outbreak to the last node. Even cutting all the edges connecting to node 8 except one, only delayed the arrival of the outbreak to Mexico City around 6 days. Reducing the coefficient D has a larger impact on the delay of the outbreak, reducing D by one order of magnitude, from 10^− 6^ to 10^− 7^, delayed the arrival of the outbreak to node 8 around 2 weeks.

The figure 2, also shows something else which is important, the time at which the first node reaches the maximum moves forward as the R0 decreases, this indicates that the isolation measures carried out in Wuhan only decreased a few tenths the R0, of the order of 5 tenths. This is because, according to the data presented by Johns Hopkins University, the report of cases begins to stabilize on approximately the in February 28, and therefore the maximum is expected to be reached close to this date.

## Discussion

This model estimates the arrival of the infectious outbreak to Mexico between March 20 and March 30. This estimation gives a time period to implement and strengthen preventive measures aimed at the general population, as well as to strengthen hospital infrastructure and training of human resources in health. However, this period should be considered as an estimate of the maximum time of arrival of the infectious outbreak to Mexico City, because the outbreak can arrive by other routes, such as sea or land by other of our borders.

This model shows that the isolation measures that can be implemented in the cities where the outbreak first arrives, although they are very important to control it locally, do not affect significantly the time to arrival to other cities. The coefficient that best controls the spread of infection in the rest of the cities, when this dispersion is by airlines, is the parameter D. Therefore, surveillance at airports should be strengthened, with special emphasis on those connecting Mexico directly or indirectly with Asian countries. Finally, this model shows that all these measures can only delay the arrival of SARS-CoV-2, but if it can be delayed long enough, it would be very important to have as much time as possible to establish the appropriate prevention and control measures. it will allow better management of the outbreak.

## Data Availability

All the data used in this work are of public access.

